# 3D Segmentation of Pathological Muscle with a Physics-Informed Latent-Regularized U-Net

**DOI:** 10.64898/2026.09.15.26363155

**Authors:** Negar Mehrabi, Nicolas C. Pégard, Geoffrey Gale Handsfield

## Abstract

**Objective:** We developed a data-efficient deep learning framework for three-dimensional segmentation of pathological musculoskeletal anatomy from magnetic resonance imaging (MRI) when only limited manual annotations are available.

**Methods:** We developed a Physics-Informed Latent-Regularized U-Net (PILR-U-Net) that combines transfer learning from healthy MRI, latent-space anatomical regularization, and elasticity-based physics-informed constraints. The physics-informed loss enforces mechanical equilibrium, near-incompressibility, and spatial smoothness of predicted deformation fields. The framework was evaluated on MRI datasets from 50 participants with cerebral palsy across 15 lower-limb musculoskeletal structures using sparse manual annotations. Performance was assessed using volumetric overlap, boundary accuracy, volume error, sensitivity, and precision. Ablation experiments evaluated the individual contributions of latent-space and physics-informed regularization.

**Results:** Our experimental results show accurate segmentations with three-dimensional Dice coefficients ranging from 0.750 to 0.943 across evaluated structures, while most structures exhibited low surface-distance errors. Predicted deformation fields maintained positive Jacobian determinants near unity and smooth strain-energy distributions. Ablation analysis showed that both regularization components improved performance, with removal of physics-informed regularization producing the largest reductions in Dice accuracy and increases in boundary error.

**Conclusion:** PILR-U-Net enables accurate and anatomically plausible segmentation of pathological musculoskeletal MRI under sparse supervision.

**Significance:** Incorporating anatomical priors and biomechanical constraints into deep segmentation networks may reduce dependence on extensive pathological annotations and support patient-specific musculoskeletal modeling and clinical analysis.

## I. Introduction

Muscle and bone segmentation from magnetic resonance images (MRI) is a valuable image analysis for musculoskeletal studies, and for the development of subject-specific models for biomechanical analysis. For example, quantitative measurements of bone and muscle shape and size are used to assess the muscular effects of pathologies such as cerebral palsy (CP) [1]. Accurate delineation of individual muscles, muscle groups, and bones from large 3D imaging datasets enables *in vivo* measurements of individual muscle and bone volumes, modeling of shape and fiber orientation, and helps understand musculoskeletal pathologies [2–4]. Manual image segmentation is highly labor-intensive, and prone to inter-observer variability. Non-AI automated segmentation routines perform poorly due to anatomical irregularities, low tissue contrast, and large morphological variability, especially in individuals with musculoskeletal pathologies [5].

Deep learning, particularly convolutional neural networks (CNNs), has shown remarkable performance in medical image segmentation tasks. Architectures such as U-Net and its derivatives have become common in biomedical image processing, providing high accuracy and spatial coherence in organ and tissue delineation [6,7]. Nevertheless, their success heavily depends on the availability of large, well-annotated datasets, which are rarely available for pathological populations [8]. As a result, purely data-driven segmentation methods overfit small training populations and fail to capture physiologically consistent muscle structures [9]. In practice, healthcare providers only segment a fraction of the available MRI data to assess the patient musculoskeletal health. Unnecessary bottlenecks reduce the clinical value of available data, and lead to imperfect diagnosis.

Recent advances in transfer learning and semi-supervised learning offer strategies to alleviate this limitation by transferring knowledge from healthy subjects to pathologies for which labeled datasets are limited [10–12]. Pretraining is performed on widely available datasets of anatomically normal MRI imaging data, allowing the model to learn generic muscle representations, then the model can be fine-tuned with smaller datasets showing pathological anatomy [10].

However, this strategy alone may still yield anatomically implausible solutions if the target pathology deviates substantially from the healthy prior. Deep networks trained solely on pixel-wise losses (Dice or cross-entropy) are agnostic to underlying anatomical constraints and may produce segmentations that are geometrically connected but anatomically erroneous [13].

To overcome these challenges, one solution is to integrate physics-based priors into deep neural architectures, leading to Physics-Informed Neural Networks (PINNs) [14]. PINNs enable networks to learn solutions that satisfy geometrical principles while fitting observed data [14,15]. In musculoskeletal modeling, physics-informed regularization can enforce equilibrium, smooth strain distribution, and near-incompressibility—properties essential for realistic muscle deformation [16]. Yet, most existing PINN applications target forward biomechanical simulations rather than image-driven segmentation and morphology mapping [17].

Latent-space regularization methods, including variational models and anatomical autoencoders, have shown promise in regularizing neural networks to produce anatomically plausible outputs [18–20]. For muscle segmentation, by encoding the manifold of healthy anatomical variability, latent regularization ensures that predicted muscle shapes remain consistent with known physiology even when the labeled pathological dataset is small [19].

Building on these insights, we developed a Physics-Informed Latent-Regularized U-Net (PILR-U-Net) (Fig. 1) that enables muscle segmentation based on deformation^1^ estimation in T1-weighted MR images of the lower limbs from adolescents with CP. Our framework combines the advantages of these three components: (1) Transfer learning from healthy muscle MRI to capture baseline anatomical knowledge [10]; (2) Latent-space anatomical regularization to enforce physiologically plausible structures during fine-tuning [18]; and (3) Physics-informed regularization which ensures anatomically consistent morphology through image slices [14]. In the first stage, we trained a residual U-Net using fully labeled healthy MRI data to learn spatial and textural representations of normal muscle anatomy [6]. We then fine-tuned the pretrained weights using a small subset of labeled slices from imaging data from one adolescent with CP, leveraging the latent prior in order to preserve anatomical plausibility [18]. Concomitantly, a physics-based loss function composed of equilibrium, divergence, and smoothness constraints regulates the predicted deformation field to satisfy equilibrium and volume preservation [16]. The resulting hybrid loss formulation couples image fidelity with anatomical plausibility and biomechanical consistency, effectively constraining the solution space to physiologically realistic muscle structures and deformation patterns, thereby enabling robust segmentation from minimal supervision.

**Figure 1.**
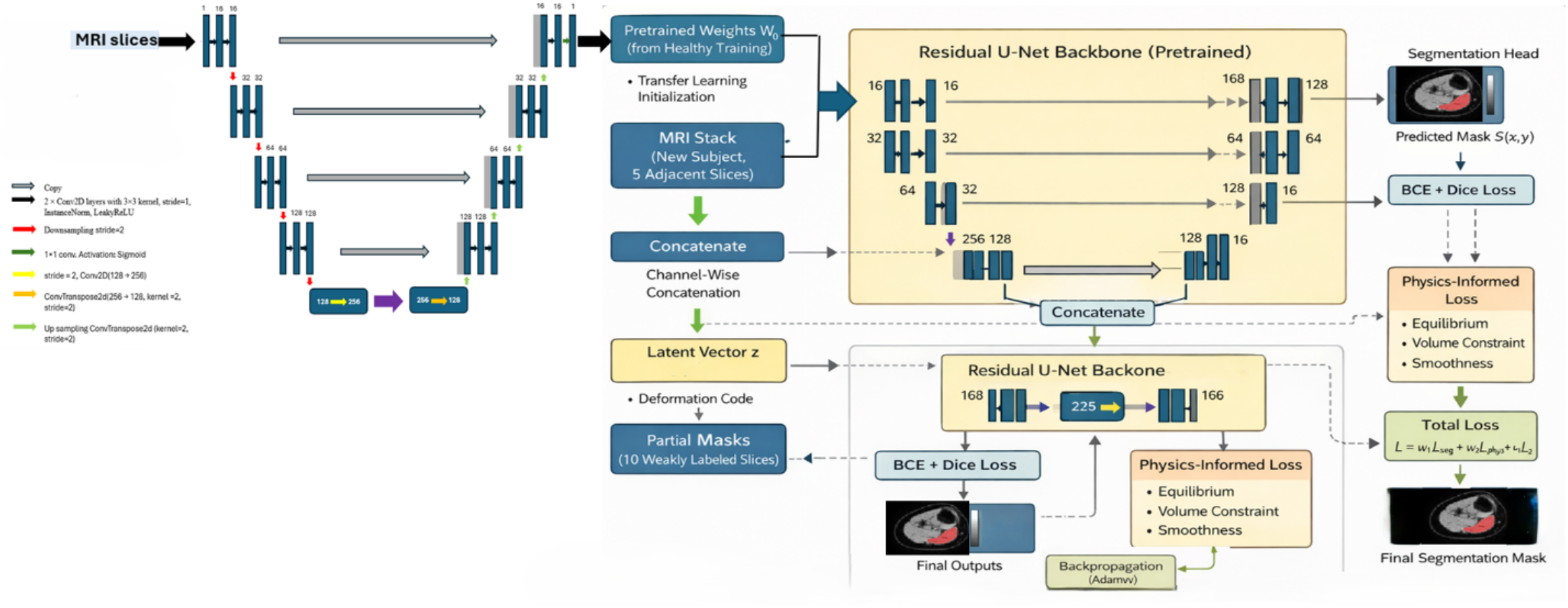
Two-stage physics-informed segmentation framework. In the first stage, a residual U-Net is trained on healthy-subject MRI data with fully annotated muscle masks to learn robust anatomical feature representations. The resulting pre-trained weights are transferred to the initialize second-stage network. In the second stage, multi-channel MRI inputs from patients with cerebral palsy, together with pre-trained feature channels, are processed through a physics-informed encoder–decoder backbone. Biomechanical constraints are enforced at the latent bottleneck space, while a dual-head architecture enables simultaneous weakly supervised segmentation and physical regularization. Partial annotations are used to guide fine-tuning, and the combined loss function integrates segmentation and physics-based terms. The framework produces anatomically consistent segmentation maps for deformed muscles in pathological conditions.

Our framework is well-suited for rare, pathological, or patient-specific musculoskeletal studies where large annotated datasets are unavailable. By integrating machine learning with anatomical priors and physics-informed constraints, this approach provides a principled framework for developing patient-specific image-based musculoskeletal models. Experimental results demonstrate that PILR U-Net improves segmentation accuracy compared to conventional U-Net architectures and generates smooth, anatomically plausible, and physically consistent deformation fields. Overall, this study advances the integration of deep learning and elasticity theory for personalized musculoskeletal analysis. It highlights how embedding physical laws and anatomical priors into neural architectures enhances model generalizability and clinical relevance, enabling data-efficient image-based diagnostics and modeling for neuro-musculoskeletal disorders such as CP.

## II. Methods

Our framework (Fig. 1), termed Physics-Informed Latent-Regularized U-Net (PILR-U-Net), aims to achieve accurate muscle segmentation in MRI from pathological populations when there is limited annotated data. The approach leverages transfer learning from healthy subjects, latent-space anatomical regularization, and physics-informed constraints derived from elasticity theory. The overall workflow (Fig. 1) consists of (1) pretraining on healthy data, (2) fine-tuning on limited CP annotations, and (3) physics-informed latent regularization during optimization.

### A. Data Acquisition and Preprocessing

#### Healthy Data

High-resolution T1-weighted MRI scans of one healthy lower limb from a previous study (21) were used for pretraining. Manual segmentation masks of major muscle groups (e.g., quadriceps, hamstrings, gastrocnemius) served as ground truth to train the base network. Each MRI volume was resampled to isotropic voxel spacing (1 mm^3^) and normalized to [0,1] intensity range.

#### Cerebral Palsy Data

MRI data from one adolescent with CP were used (Fig. 2) for fine-tuning. Only a limited number of manually labeled slices (10) were used. The remaining unlabeled slices were used via semi-supervised regularization. All data were aligned to a common anatomical frame using affine registration and cropped to muscle regions of interest.

**Figure 2.**
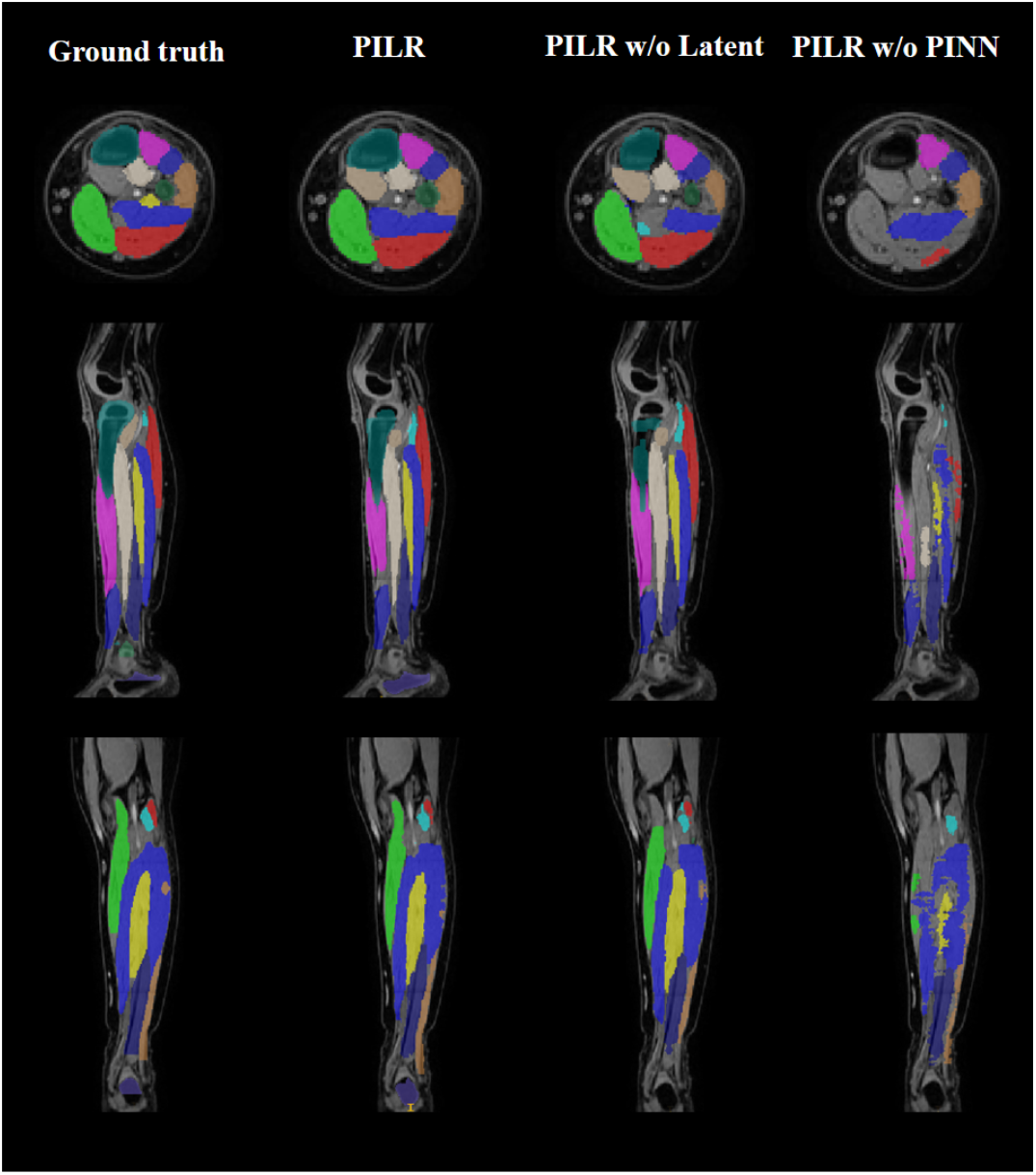
Ablation study comparing the complete PILR-U-Net with two reduced variants: PILR-U-Net (w/o Latent Regularization) and PILR-U-Net (w/o Physics-Informed Regularization) for four different subjects. The model demonstrates close anatomical agreement with expert references, accurately preserving muscle boundaries and structural details.

**Figure 3.**
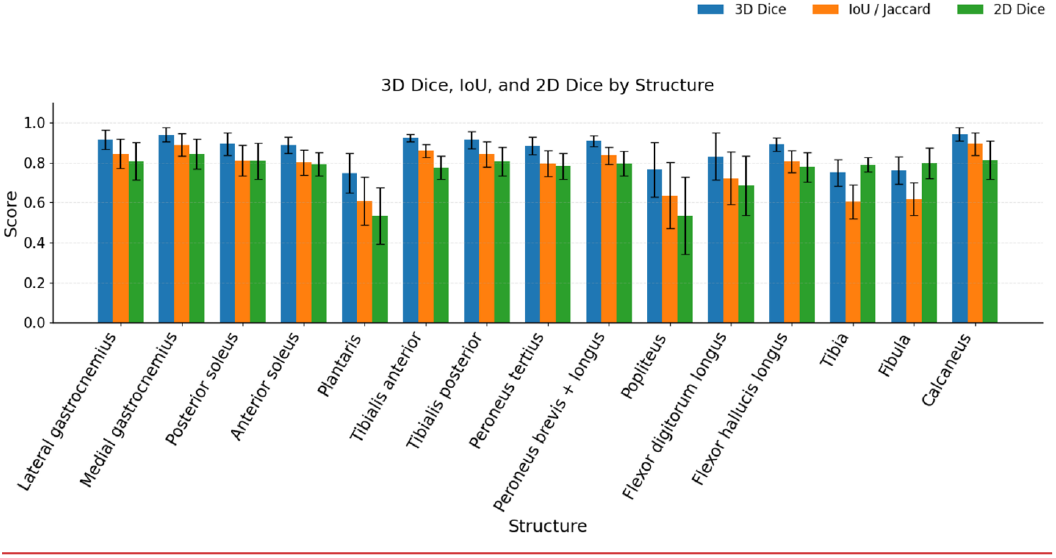
Segmentation accuracy (overlap-based metrics) Comparison of segmentation accuracy across all anatomical structures using 3D Dice, 2D Dice, and Intersection-over-Union (IoU). High Dice and IoU values across most muscles demonstrate strong spatial agreement between predicted and ground truth segmentations. Error bars represent mean ± standard deviation, indicating consistent performance despite limited annotated data.

### B. Network Architecture

The segmentation network is a Residual U-Net composed of encoder–decoder pathways with skip connections and bottleneck residual blocks to preserve gradient flow and capture multi-scale features (Fig. 1). Each encoder stage applies two 3×3 convolutions followed by batch normalization and ReLU activation, while residual links connect adjacent feature maps to improve stability. The decoder mirrors the encoder with transposed convolutions for upsampling.

A latent-space regularization module is embedded between the bottleneck and decoder. This module introduces a variational latent encoder that learns an anatomical prior distribution *p*_*θ*_ (*z*∣*X*_*healthy*_) from pre-trained healthy data. During fine-tuning on CP data, the latent posterior *q*_*Φ*_ (*z*|*X*_*CP*_) is regularized by minimizing the Kullback–Leibler divergence:

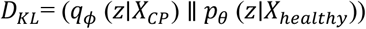

which enforces statistical proximity between pathological and healthy latent representations. This constraint promotes physiologically plausible muscle morphology under pathological conditions.

### C. Physics-Informed Regularization

To constrain the learning process with anatomical realism, the network incorporates a Physics-Informed Regularization (PIR) module derived from the equilibrium laws of linear elasticity. The predicted segmentation boundaries implicitly encode a displacement field **u**(**x**) which represents a deformation between the reference (healthy) and patient (CP) configurations. From this displacement, local strain, stress, and equilibrium residuals can be estimated and used as differentiable penalties in the loss function.

The general equilibrium equation is expressed as ∇⋅σ+f=0, where **σ**is the Cauchy stress tensor, **f** denotes body forces (neglected here, **f** = 0), and **σ**=**C**:**ε, ε**=1/2(∇**u**+(∇**u**)**T**) are the constitutive and strain relations with **C** being the isotropic elastic tensor.

#### (a) Equilibrium Loss — ℒ_phys_

This term enforces static force balance throughout the predicted deformation field:

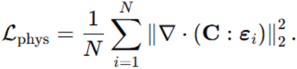

It penalizes residual stresses that would otherwise create unreasonable internal forces. Minimizing ℒ_phys_ encourages the network to produce displacement fields that satisfy mechanical equilibrium, stabilizing regions with large intensity gradients where pure data losses might overfit.

#### (b) Divergence Loss — ℒ_div_

To avoid unrealistic local volume changes (compression or dilation) within soft muscle tissue, a divergence constraint is added:

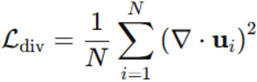

This term reflects the nearly incompressible property of skeletal muscle, which is regarded as nearly isochoric. Reducing ℒ_div_ ensures that the predicted deformation preserves local volume and prevents fold-overs in the mapping.

#### (c) Smoothness Loss — ℒ_smooth_

Spatial continuity of displacement is imposed through a first-order gradient penalty:

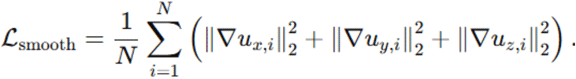

This encourages gradual spatial variation and eliminates spurious oscillations or noise in the displacement field. It acts similarly to Tikhonov regularization on deformation gradients, promoting smooth strain distributions across muscle boundaries.

#### (d) Combined Physics-Informed Loss

The overall physics term incorporated into the total objective is

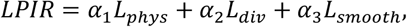

where coefficients *α*_1_, *α*_2_, *α*_3_ control the relative strength of each constraint.

In practice, these were empirically set to maintain numerical stability (*α*_1_ = 1.0, *α*_2_ = 0.5, *α*_2_ = 0.1).

By minimizing ℒ_PIR_ jointly with the segmentation and latent regularization losses, the network is guided toward deformation fields that (1) satisfy mechanical equilibrium, (2) preserve tissue volume, and (3) remain spatially smooth. This integration of continuum-mechanical priors from elasticity theory effectively suppresses anatomically implausible segmentations of the predicted CP muscle deformations.

### D. Multi-Objective Loss Function

The total loss function ℒ_*total*_was formulated as a weighted combination of segmentation, latent, and physics-based components:

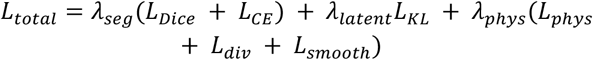

where:

ℒ_*Dice*_ and ℒ_*CE*_ ensure accurate muscle segmentation, ℒ_*KL*_ enforces latent anatomical consistency.

The weighting factors *λ*_*seg*_, *λ*_*latent*_, *λ*_*phys*_ were empirically tuned to balance segmentation accuracy.

### E. Training and Fine-Tuning Strategy

The training process comprised two main stages: (1) Pretraining on Healthy Data: The Residual U-Net was trained on fully labeled healthy muscle MRI data for 100 epochs using the AdamW optimizer (learning rate = 1e-4, batch size = 8). The model weights were saved and used as the initialization for fine-tuning. (2) Fine-Tuning on CP Data: The pretrained model was fine-tuned using the limited labeled slices from the CP dataset. Unlabeled slices were included via semi-supervised learning using the physics and latent regularization terms. Fine-tuning employed a smaller learning rate (1e-5) and dynamic early stopping based on validation loss stabilization.

### F. Implementation Details

All experiments were conducted in PyTorch 2.2.0 using an NVIDIA RTX GPU (24 GB). Data augmentation (rotation, elastic deformation, and Gaussian noise) was applied to enhance generalization. The total training time for both stages was approximately 7 hours. Model checkpoints and optimizer states were saved to ensure reproducibility.

### G. Evaluation Metrics

The proposed Physics-Informed Latent-Regularized U-Net (PILR-U-Net) framework was evaluated on MRI datasets from 50 participants with cerebral palsy (CP) obtained from Bolsterlee et al. (21), comprising annotated lower-limb muscles and bones. For each participant, only 10 randomly selected manually segmented slices were provided as sparse labeled supervision, while the complete corresponding MRI stack was used as image input. Using these limited annotations together with the full MRI volume, PILR-U-Net generated complete three-dimensional segmentations of the evaluated lower-limb muscles and bones. The resulting predictions were compared with the complete reference segmentations provided by Bolsterlee et al. Segmentation performance was quantitatively evaluated using multiple complementary metrics assessing volumetric overlap, boundary accuracy, classification performance, and geometric consistency, as summarized in (See Supplementary Table 1) and (See Supplementary Fig. 1– 3).

Dice Similarity Coefficient (DSC) was employed to measure the degree of spatial overlap between the predicted segmentation and the ground truth. DSC is defined as twice the intersection of the predicted and reference regions divided by the sum of their volumes and ranges from 0 (no overlap) to 1 (perfect agreement). Boundary accuracy was assessed using the 95th percentile Hausdorff Distance (HD95), which quantifies the maximum surface-to-surface deviation between the predicted and reference segmentations while reducing sensitivity to outliers. Distances were computed in physical space using voxel spacing and are reported in millimeters. Average Symmetric Surface Distance (ASSD) was calculated to measure the mean bidirectional distance between corresponding surface points of the predicted and ground truth segmentations. ASSD provides an estimate of the average boundary deviation and reflects overall contour accuracy.

In addition to overlap- and surface-based metrics, volumetric consistency was evaluated by computing the relative volume error, defined as the percentage difference between the predicted and reference muscle volumes. Sensitivity (recall) and precision were also computed to assess classification behavior, reflecting the ability of the proposed method to correctly identify true muscle tissue and avoid false positive predictions, respectively. Computed deformation fields were assessed for reasonableness by analyzing the strain energy density and Jacobian determinant distributions derived from the estimated deformation fields. These measures were used to confirm the absence of non-physical distortions such as folding or unrealistic volumetric compression and expansion. All evaluation metrics were computed on a per-subject basis for each target muscle. Quantitative results from all 50 test subjects were aggregated, and the mean and standard deviation were calculated for each metric and each muscle. These summary statistics were used to assess the accuracy, robustness, and inter-subject consistency of the proposed segmentation framework.

### H. Ablation Study Design

To quantify the individual contributions of the two principal components of the proposed framework, namely latent-space anatomical regularization and physics-informed regularization, an ablation study was conducted. Two additional model variants were derived from the complete PILR-U-Net architecture by independently removing one regularization component while preserving all remaining architectural, optimization, and training settings. This design enabled the isolated evaluation of each component while ensuring a fair comparison among all model variants.

The first ablation model excluded the latent-space anatomical regularization module while retaining healthy-data pre-training, the physics-informed regularization module, the network architecture, optimization strategy, and all other loss terms. Consequently, the Kullback–Leibler divergence loss (ℒ_*KL*_) was removed from the optimization objective, whereas the segmentation and physics-informed loss components remained unchanged. The second ablation model excluded the physics-informed regularization module while preserving latent-space anatomical regularization, healthy-data pre-training, network architecture, and all remaining training procedures. In this configuration, the equilibrium (*L*_*phys*_), divergence (*L*_*div*_), and smoothness (*L*_*smooth*_) loss terms were omitted from the objective function, while the segmentation and latent-space regularization losses were retained. All three models—the complete PILR-U-Net, the model without latent-space regularization, and the model without physics-informed regularization—were trained and evaluated using identical preprocessing procedures, data augmentation strategies, optimization parameters, learning schedules, and evaluation metrics. Segmentation performance was assessed using the Dice Similarity Coefficient (DSC), Jaccard Index, 95th percentile Hausdorff Distance (HD95), Average Symmetric Surface Distance (ASSD), relative volume error, sensitivity, and precision. This experimental design ensured that any observed differences in performance could be attributed exclusively to the inclusion or exclusion of the corresponding regularization component.

The objective of the ablation study was to determine the individual contribution of latent-space anatomical regularization and physics-informed regularization to segmentation accuracy, boundary delineation, volumetric preservation, and anatomical consistency. Owing to the substantial computational cost associated with retraining multiple deep learning models under identical experimental conditions, the ablation experiments were conducted using the same 10 participants across all three model variants. All statistical analyses were subsequently performed using these paired participant-level segmentation results.

### I. Statistical Analysis

Statistical analysis was performed to evaluate differences in segmentation performance among the complete PILR-U-Net and the two ablation models. For each musculoskeletal structure, paired performance measurements obtained from the same 10 participants were compared across the three model variants.

Overall differences among the three models were first evaluated using the non-parametric Friedman test, which ranks the performance of each model within each participant to determine whether statistically significant differences exist among the three paired model configurations. When appropriate, pairwise comparisons between the complete PILR-U-Net and each ablation model, as well as between the two ablation models, were subsequently performed using the Wilcoxon signed-rank test based on paired participant-level differences.

For both the Friedman and Wilcoxon tests, raw p-values were calculated from the corresponding null distributions of the test statistics. To account for multiple statistical comparisons across the 15 evaluated musculoskeletal structures, p-values within each family of tests were adjusted using the Benjamini– Hochberg false discovery rate (FDR) correction. Statistical significance was defined as an FDR-adjusted p < 0.05.

All statistical analyses were performed using Python 3.11 with the SciPy and statsmodels libraries.

## III. Results

### A. Overall Segmentation Performance

Comparison of the predicted segmentations with the manually annotated reference masks from Bolsterlee et al. (21) demonstrated that the proposed PILR-U-Net achieved consistently high segmentation fidelity across all evaluated musculoskeletal structures. The 3D Dice coefficient (See Supplementary Table 1) (See Supplementary Fig. 1), a measure of volumetric agreement, ranged from 0.7497 ± 0.0674 (tibia) to 0.9427 ± 0.0331 (calcaneus), highlighting robust overlap with reference segmentations. Similarly, the Jaccard index varied between 0.6041 ± 0.0854 (tibia) and 0.8933 ± 0.0572 (calcaneus). Slice-wise 2D Dice scores were consistently high, ranging from 0.5342 ± 0.1925 (popliteus) to 0.8432 ± 0.0756 (medial gastrocnemius), which demonstrated stable segmentation performance along the longitudinal axis of the muscles. Muscles and bones with simple morphology and high tissue contrast, including medial gastrocnemius, tibialis anterior, and calcaneus, achieved the highest volumetric accuracy (3D Dice ≥ 0.92). In contrast, elongated or low-contrast muscles and bones, such as plantaris, popliteus, tibia, and fibula, exhibited moderately reduced performance (3D Dice ≤ 0.76), although still well within clinically acceptable boundaries.

### B. Boundary Accuracy

Boundary delineation (See Supplementary Table 1) (See Supplementary Fig. 2) was quantified using the 95th percentile Hausdorff Distance (HD95) and the Average Symmetric Surface Distance (ASSD). HD95 ranged from 2.446 ± 2.218 mm (calcaneus) to 39.615 ± 54.048 mm (popliteus). The majority of muscles and bones exhibited HD95 < 6 mm, indicating acceptably low maximum surface deviations. Elevated HD95 values in plantaris, popliteus, tibia, and fibula reflect more intricate surface geometries and lower MRI contrast, consistent with challenges in delineating these structures. ASSD values were consistently low across the cohort, ranging from 0.591 ± 0.358 mm (Calcaneus) to 7.017 ± 10.206 mm (Popliteus), confirming high average boundary fidelity and accurate contouring of muscle surfaces even in complex regions. Collectively, these findings demonstrate high boundary delineation accuracy and robust surface agreement across anatomically diverse musculoskeletal structures.

### C. Volumetric Consistency

Volumetric accuracy (See Supplementary Table 1) (See Supplementary Fig. 3), assessed via relative volume error, remained tightly constrained around zero, ranging from -0.3344 ± 0.1204 (tibia) to 0.2158 ± 0.6023 (popliteus). These results indicate that the proposed framework preserves anatomical volume with minimal systematic over- or under-segmentation. Muscles and bones exhibiting higher boundary variability, such as plantaris, popliteus, flexor hallucis longus, and tibia, displayed larger standard deviations, reflecting inter-subject anatomical variability and segmentation complexity, yet still maintained clinically acceptable volumetric fidelity.

### D. Classification Performance

We used sensitivity (recall) and precision (See Supplementary Table 1) (Supplementary Fig. 6) to assess correct identification of muscle tissue within images. Sensitivity ranged from 0.6275 ± 0.0947 (tibia) to 0.9311 ± 0.0432 (calcaneus), indicating robust detection of true muscle/bone voxels. Precision ranged from 0.7376 ± 0.1481 (plantaris) to 0.9566 ± 0.0451 (calcaneus), reflecting effective suppression of false positives. Muscles with lower sensitivity or precision (plantaris, popliteus, tibia, and fibula) were those with complex morphology, low tissue contrast, or proximity to adjacent structures, consistent with volumetric and boundary accuracy observations.

### E. Muscle-Specific Performance

Analysis of individual muscle metrics (See Supplementary Table 1) revealed distinct performance trends. High-performing musculoskeletal structures: medial gastrocnemius, tibialis anterior, peroneus longus, and calcaneus achieved 3D Dice > 0.90, HD95 < 5 mm, ASSD < 1 mm, and high sensitivity and precision (>0.90). These muscles/bones are anatomically compact, well-defined, and high-contrast, facilitating robust segmentation. Moderate-performing muscles: lateral gastrocnemius, posterior soleus, anterior soleus, tibialis posterior, peroneus tertius, and flexor digitorum longus demonstrated 3D Dice between 0.88–0.91, HD95 up to 10.8 mm, and volume errors within ±0.11, reflecting minor boundary complexity while maintaining reliable segmentation accuracy. Challenging muscles: plantaris, popliteus, tibia, and fibula achieved lower Dice scores (2D Dice: 0.534–0.761, 3D Dice: 0.749–0.765) and higher boundary deviations (HD95 up to 39.6 mm, ASSD up to 7 mm), primarily due to elongated shapes, low tissue contrast, or adjacency to other structures.

### F. Deformation field assessment

Jacobian determinants and strain energy density distributions (See Supplementary Table 1) were derived from the predicted deformation fields and used to evaluate the biomechanical plausibility of the predicted deformation fields. Across all muscles, Jacobian determinants remained positive and near unity, while strain energy density distributions were smooth, confirming the absence of non-physical distortions.

### G. Ablation Study

To quantify the individual contributions of latent-space anatomical regularization and physics-informed regularization, an ablation study was conducted by systematically removing each component while maintaining identical network architecture, training procedures, preprocessing, optimization settings, and evaluation metrics. Quantitative performance for the complete PILR-U-Net framework and the two ablation models is summarized in (See Supplementary Tables 2 and 3), while (See Supplementary Fig.s 4 and 5) provide a visual comparison of the corresponding 3D Dice coefficients and HD95 values.

Removal of the latent-space regularization module resulted in a modest but consistent reduction in segmentation performance across the majority of muscles and bones. Although overall segmentation accuracy remained acceptable, decreases were observed in volumetric overlap metrics, including the Dice Similarity Coefficient and Jaccard Index, together with moderate increases in boundary-based metrics (HD95 and ASSD). These observations indicate that latent-space anatomical regularization improves segmentation robustness by encouraging pathological muscle representations to remain close to anatomically plausible latent distributions learned from healthy subjects.

The effect of removing the physics-informed regularization module was considerably more pronounced. Across nearly all evaluated structures, Dice coefficients decreased substantially, while HD95 and ASSD increased markedly. Several muscles that achieved Dice coefficients exceeding 0.90 using the complete PILR-U-Net exhibited reductions to approximately 0.65–0.80 following removal of the physics-informed regularization. Likewise, HD95 increased substantially in anatomically challenging structures, including the plantaris, popliteus, tibia, and fibula, indicating poorer boundary localization and reduced anatomical consistency.

Overall, comparison of the three model variants demonstrates that both latent-space anatomical regularization and physics-informed regularization contribute positively to segmentation performance. However, removal of the physics-informed regularization consistently produced the largest deterioration in both overlap- and boundary-based metrics, indicating that biomechanical constraints contributed substantially more to segmentation accuracy and anatomically realistic boundary delineation than latent-space regularization alone. The complete PILR-U-Net, incorporating both regularization strategies, consistently achieved the highest overall performance.

The statistical significance of the observed differences among the three model variants was evaluated using the Friedman test followed by pairwise Wilcoxon signed-rank tests with Benjamini–Hochberg false discovery rate (FDR) correction (See Supplementary Tables 4 and 5).

For the **3D Dice coefficient** (See Supplementary Table 4), the Friedman test demonstrated significant overall differences among the three model variants for the majority of evaluated musculoskeletal structures following FDR correction. Pairwise comparisons further showed that the complete PILR-U-Net significantly outperformed the model without physics-informed regularization for most muscles and bones (*FDR-corrected p* < 0.05). In contrast, comparisons between the complete model and the model without latent-space regularization generally did not reach statistical significance after FDR correction, although a consistent reduction in segmentation accuracy was observed across many structures. These findings suggest that latent-space regularization provided complementary improvements in segmentation robustness, whereas physics-informed regularization contributed substantially more to overall segmentation accuracy. Similarly, for **HD95** (See Supplementary Table 5), statistically significant overall differences were identified for several musculoskeletal structures after FDR correction. Pairwise comparisons again demonstrated significantly lower HD95 values for the complete PILR-U-Net relative to the model without physics-informed regularization, indicating improved boundary delineation and anatomical consistency. Differences between the complete model and the latent-space ablation model were generally smaller and were not statistically significant for most structures after FDR correction.

Collectively, the quantitative and statistical analyses demonstrated that physics-informed regularization contributed the largest improvement in segmentation accuracy and boundary delineation, while latent-space regularization provided complementary gains in anatomical robustness and representation learning. These findings demonstrate that the integration of latent-space anatomical priors and physics-informed regularization is necessary to achieve optimal segmentation performance under limited pathological supervision.

## IV. Discussion

Accurate segmentation of skeletal muscles and bones in pathological populations remains a major challenge in medical imaging applications for orthopaedics, biomechanics, sports medicine, and musculoskeletal clinical applications [22,23]. Traditional deep learning-based segmentation methods rely heavily on large, annotated datasets and often fail when confronted with atrophied or atypical muscle morphologies, as observed in cerebral palsy (CP) and other neuromuscular disorders [24–26]. The present study introduces a Physics-Informed Latent-Regularized U-Net (PILR-U-Net) that successfully addresses these limitations by integrating three complementary strategies: transfer learning from healthy datasets, latent-space anatomical regularization, and physics-informed constraints [27–30]. Across 50 subjects and 15 structures, our framework demonstrated robust volumetric, surface, and classification accuracy, while preserving anatomical plausibility and enabling physiologically interpretable deformation mapping.

### A. BRIDGING Data Scarcity and Anatomical Plausibility

One of the critical challenges in pathological muscle imaging is the scarcity of labeled data. While fully supervised networks, including U-Net and its derivatives, achieve high segmentation accuracy in healthy populations, their performance deteriorates sharply in pathological cases due to anatomical deviations and low tissue contrast [31–35]. PILR-U-Net addresses this by leveraging transfer learning from healthy muscle MRI [27]. Pre-training on healthy datasets allows the network to capture baseline anatomical and structural representations, which are subsequently fine-tuned to the pathological domain using only a limited subset of labeled CP slices. This approach minimizes overfitting while ensuring that the network retains an internal representation of physiologically plausible muscle structures, a critical advantage over conventional fully supervised or semi-supervised frameworks [27–29].

Latent-space regularization further reinforces anatomical plausibility by constraining predictions to a learned manifold of healthy variability [36–38]. Unlike previous methods that rely solely on pixel-wise or voxel-wise losses, latent priors encode the spatial and morphological relationships inherent in healthy anatomy, guiding the network to produce predictions that are both anatomically consistent and adaptive to pathological deviations.

### B. Physics-Informed Regularization for Anatomical Consistency

Latent priors do not inherently enforce anatomical fidelity. To address this, PILR-U-Net integrates physics-inspired loss functions motivated by linear elasticity, including equilibrium, divergence, and smoothness constraints [40–42]. These terms are designed to encourage predicted deformation fields to satisfy mechanical principles such as force balance, near-incompressibility, and spatial continuity [41]. As a result, the network produces smooth displacement and strain fields, promoting anatomically plausible outputs consistent with diffeomorphic deformation models [39,40–42]. Furthermore, the ablation analysis demonstrated that removal of the physics-informed regularization resulted in the largest performance degradation observed in this study, with substantial increases in HD95 and ASSD values across multiple structures. This finding provides quantitative evidence that the equilibrium, divergence, and smoothness constraints contribute directly to segmentation robustness and anatomical consistency rather than acting solely as auxiliary regularization terms.

The efficacy of this approach is supported by consistently positive Jacobian determinants near unity across all muscles, indicating the absence of folding, unrealistic compression, or expansion [39]. Similarly, smooth strain energy density distributions suggest that the predicted deformation fields are mechanically coherent. This represents a potential improvement over traditional CNN-based methods, which may produce anatomically inconsistent segmentations [43].

### C. Ablation Analysis of Latent and Physics-Informed Regularization

The ablation study provides direct evidence for the individual and complementary contributions of latent-space anatomical regularization and physics-informed regularization within the proposed PILR-U-Net framework. Two modified variants of the proposed architecture were evaluated: (1) a model without latent-space regularization and (2) a model without physics-informed regularization, while preserving identical network architecture, pre-training strategy, preprocessing, optimization parameters, and training protocol. This experimental design ensured that performance differences could be attributed solely to the removal of the corresponding regularization component. Removal of the latent-space regularization resulted in a moderate but consistent reduction in segmentation performance across the majority of evaluated muscles and bones. Although the overall Dice Similarity Coefficient remained relatively high for many structures (Supplementary Fig. 7), decreases in volumetric overlap were accompanied by modest increases in HD95 (Supplementary Fig. 8) and ASSD. These findings suggest that latent-space anatomical regularization contributes primarily by encouraging pathological muscle representations to remain close to anatomically plausible latent distributions learned from healthy subjects. Consequently, the network becomes less susceptible to overfitting when only limited pathological annotations are available and produces more anatomically consistent segmentations, particularly for structures exhibiting greater morphological variability.

In contrast, removal of the physics-informed regularization produced substantially larger performance degradation across nearly all evaluated structures. Dice Similarity Coefficients decreased considerably, while HD95, ASSD, and relative volume error increased, indicating deterioration in both volumetric agreement and boundary localization. The largest reductions were consistently observed in anatomically challenging structures, including the plantaris, popliteus, tibia, and fibula, where low tissue contrast, elongated morphology, and close proximity to adjacent anatomical structures make accurate segmentation particularly difficult. These observations indicate that the equilibrium, divergence, and smoothness constraints effectively suppress anatomically implausible predictions, stabilize boundary estimation, and improve the geometric consistency of the predicted segmentations.

The statistical analysis further strengthens these observations. Friedman tests demonstrated significant overall differences among the three model variants for the majority of evaluated musculoskeletal structures. Subsequent pairwise Wilcoxon signed-rank tests with Benjamini–Hochberg false discovery rate (FDR) correction showed that the complete PILR-U-Net significantly outperformed the model without physics-informed regularization for most structures in terms of both segmentation accuracy (3D Dice) and boundary accuracy (HD95). In comparison, removal of the latent-space regularization generally produced smaller reductions in performance, and fewer pairwise comparisons remained statistically significant after FDR correction. These statistically validated findings indicate that the largest measurable improvement in segmentation performance is attributable to the physics-informed regularization, whereas latent-space regularization provides complementary improvements in anatomical robustness and generalization under limited supervision.

Importantly, the two regularization strategies appear to fulfill complementary roles within the proposed framework. The latent-space regularization primarily improves representation learning by embedding pathological anatomy within a physiologically meaningful latent manifold learned from healthy subjects, thereby enhancing generalization under sparse supervision. Conversely, the physics-informed regularization directly constrains the optimization process through biomechanical principles, encouraging mechanically consistent deformation fields and anatomically realistic segmentation boundaries. The superior performance of the complete PILR-U-Net, together with the statistical significance demonstrated by the ablation analysis, indicates that these two mechanisms act synergistically rather than redundantly. Integrating both anatomical prior knowledge and biomechanical constraints enables the framework to achieve higher segmentation accuracy, improved boundary fidelity, and greater anatomical plausibility than either strategy alone.

### D. Muscle-Specific Performance and Anatomical Considerations

Analysis of individual muscles and bones revealed predictable variations in segmentation performance, correlating with anatomical complexity and imaging characteristics [32,34]. High-performing muscles and bones, such as medial gastrocnemius, tibialis anterior, peroneus longus, and calcaneus, exhibited 3D Dice coefficients >0.90, HD95 <5 mm, and ASSD <1 mm. These muscles are relatively compact, well-defined, and present high MRI contrast, facilitating accurate segmentation. Conversely, elongated or low-contrast muscles/bones, including plantaris, popliteus, tibia, and fibula, demonstrated moderately reduced performance (3D Dice 0.749–0.765; HD95 up to 39.6 mm), reflecting inherent challenges associated with complex morphology, proximity to adjacent structures, and tissue heterogeneity.

Standard U-Net or V-Net architectures trained on healthy datasets typically suffer from Dice reductions exceeding 0.15–0.20 in pathological muscles [31–35], while semi-supervised or transfer learning approaches often lack explicit anatomical constraints, resulting in unrealistic deformation fields [27–29]. By combining anatomical priors with physics-informed regularization, PILR-U-Net overcomes some of these limitations, offering possibilities for rapid segmentation of rare or pathological data even without specific training datasets.

### E. Comparative Analysis with Prior Work

Recent studies on muscle and musculoskeletal MRI segmentation can be broadly categorized into three approaches: fully supervised CNN-based methods, semi-supervised or transfer learning strategies, and physics-informed neural networks (PINNs). Fully supervised CNN models typically rely on large-scale annotated datasets, often requiring tens to hundreds of subjects to achieve robust performance in muscle MRI segmentation, as demonstrated in lumbar paraspinal segmentation work using 76 participants (47). Semi-supervised and transfer learning methods have been introduced to mitigate annotation scarcity by leveraging unlabeled data or pre-trained models. For example, semi-supervised learning with only ∼10% of labeled slices has shown segmentation performance comparable to fully supervised training (48). However, many semi-supervised learning frameworks focus on leveraging unlabeled data without explicit anatomical constraints and may still exhibit reduced structural coherence compared with fully labeled models (49). Physics-informed neural networks have primarily been applied to forward biomechanical modeling tasks, such as integrating finite element analysis with PINNs for tissue property prediction in spine biomechanics (50) and predicting muscle forces and joint kinematics under embedded physical laws (51). Their application to image segmentation is limited, underscoring the novelty of the current integration of elasticity-based constraints within a segmentation framework. In contrast to previous studies, the proposed framework simultaneously integrates transfer learning, latent-space anatomical regularization, and physics-informed regularization within a single segmentation architecture. Furthermore, the present study quantitatively demonstrates the individual contribution of each component through an ablation study with statistical validation, providing evidence that the combined framework offers complementary advantages over either regularization strategy alone.

### F. Limitations and Future Directions

Several limitations warrant consideration in this study. First, the current work focuses on a single pathological population (cerebral palsy), and validation on larger cohorts and additional neuromuscular disorders is important to establish broader generalizability [52]. Second, the physics-informed regularization adopted in this study is based on linear elasticity assumptions, which approximate muscle tissue as homogeneous and linearly deformable. While this formulation ensures numerical stability, future work could incorporate nonlinear, anisotropic, or patient-specific constitutive models derived from electrography or diffusion MRI to better capture large deformations, regional stiffness variations, and fiber-dependent mechanics [53]. Third, the latent anatomical prior was learned from a limited number of healthy subjects; expanding the training set could capture a broader spectrum of anatomical variability, including population- and age-related differences [54]. Importantly, because the proposed framework requires only minimal pathological annotations, the availability of larger healthy datasets would not diminish its time-saving advantages in clinical applications. Finally, computational efficiency remains a consideration for real-time clinical deployment, and future work will explore model optimization and acceleration strategies to facilitate integration into routine clinical workflows [55].

#### 4.7. Clinical Implications and Translational Relevance

Accurate segmentation is useful for quantifying muscle atrophy, surgical planning, and evaluating functional outcomes in neuromuscular disorders such as CP [44–46]. The data-efficient nature of the framework addresses a major bottleneck in clinical translation: the limited availability of labeled pathological datasets. By producing anatomically plausible predictions from limited training data, this framework may be useful in the future for patient-specific musculoskeletal simulations and may prove useful for tasks requiring intensive segmentation such as longitudinal monitoring and personalized therapy planning. We hope that this improved and data-efficient pipeline may facilitate broader adoption in clinical workflows and rare disease studies. While the accuracy of the current method varies across muscles and bones, this accuracy may be improved over time through muscle-specific tailoring, but further research is warranted. There may be additional applications in clinical programs for longitudinal monitoring and personalized therapy planning.

## V. Conclusion

In conclusion, this study presents PILR-U-Net, a novel segmentation framework that integrates transfer learning, latent-space anatomical regularization, and physics-informed regularization within a unified deep learning architecture for skeletal muscle and bone segmentation under limited annotated pathological data. Across 50 participants and 15 musculoskeletal structures, the proposed framework achieved robust volumetric overlap, accurate boundary delineation, and anatomically consistent segmentations, demonstrating its ability to generalize across structures with diverse morphology and imaging characteristics. The ablation study, supported by statistical analysis using Friedman tests, pairwise Wilcoxon signed-rank tests, and Benjamini–Hochberg false discovery rate (FDR) correction, demonstrated that both latent-space anatomical regularization and physics-informed regularization contribute to the overall performance of the framework. While latent-space regularization improved representation learning and promoted anatomically plausible segmentations under sparse supervision, physics-informed regularization produced the largest and most statistically significant improvements in segmentation accuracy, boundary fidelity, and geometric consistency. These findings demonstrate that the two regularization strategies provide complementary benefits, with their combined integration consistently outperforming either component individually.

By combining anatomical prior knowledge with biomechanical constraints, PILR-U-Net addresses two major challenges in musculoskeletal image segmentation: limited availability of annotated pathological datasets and the generation of anatomically realistic segmentations. The proposed framework therefore provides a robust foundation for future image-driven musculoskeletal analysis, patient-specific biomechanical modeling, digital twin development, and AI-assisted clinical decision support in both healthy and pathological populations [36–38,39–41].

Ethics statement: This study involved secondary analysis of previously collected, de-identified MRI data obtained from Bolsterlee et al. [21] with permission from the original investigators. No new participants were recruited, and the investigators had no access to identifiable private information. The original data collection was conducted under the institutional ethics approval and informed-consent procedures described in the original publication [21]. The present secondary analysis was determined not to constitute human subjects research and therefore did not require additional IRB approval.

## Data Availability

The MRI data analyzed in this study were previously collected and were obtained from Bolsterlee et al. with permission from the original investigators. The data were de-identified prior to analysis. Access to the underlying imaging data is subject to the policies and permissions of the original data custodians

## Footnotes

1 Here, deformation refers to shape differences between a typical healthy muscle and a pathological muscle.

