## Supplementary Document for "3D Segmentation of Pathological Muscle with a Physics-Informed Latent-Regularized U-Net"

### Supplementary material

Negar Mehrabi, Nicolas C. Pégard, and Geoffrey Gale Handsfield

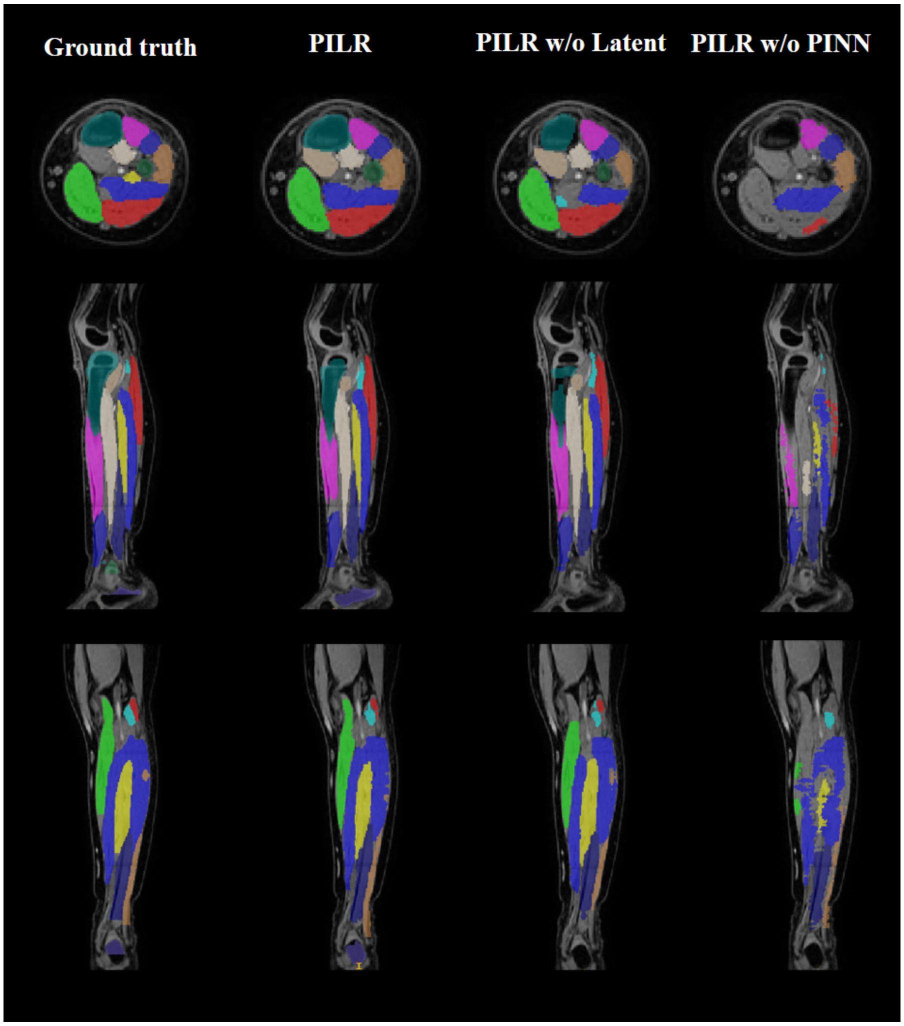


**Supplementary Figure (1).** Ablation study comparing the complete **PILR-U-Net** with two reduced variants: **PILR-U-Net (w/o Latent Regularization)** and **PILR-U-Net (w/o Physics-Informed Regularization) for four different subjects**. The model demonstrates close anatomical agreement with expert references, accurately preserving muscle boundaries and structural details.

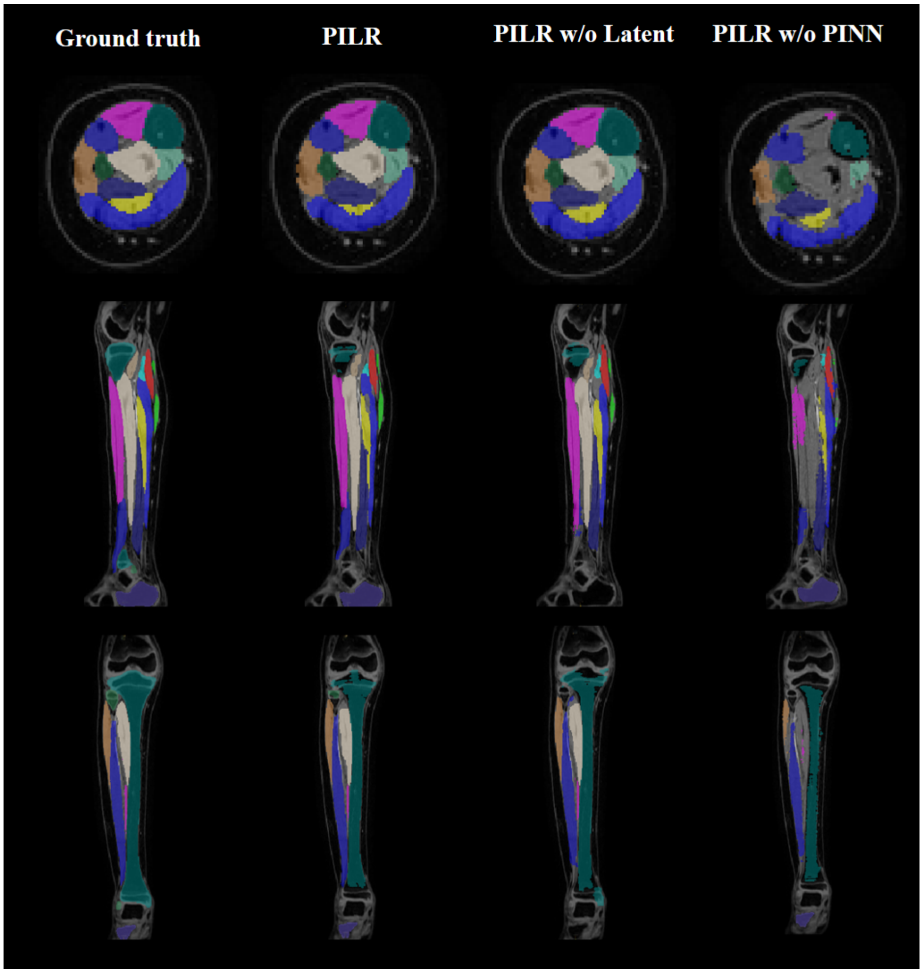


**Supplementary Figure (2).** Ablation study comparing the complete **PILR-U-Net** with two reduced variants: **PILR-U-Net (w/o Latent Regularization)** and **PILR-U-Net (w/o Physics-Informed Regularization) for four different subjects**. The model demonstrates close anatomical agreement with expert references, accurately preserving muscle boundaries and structural details.

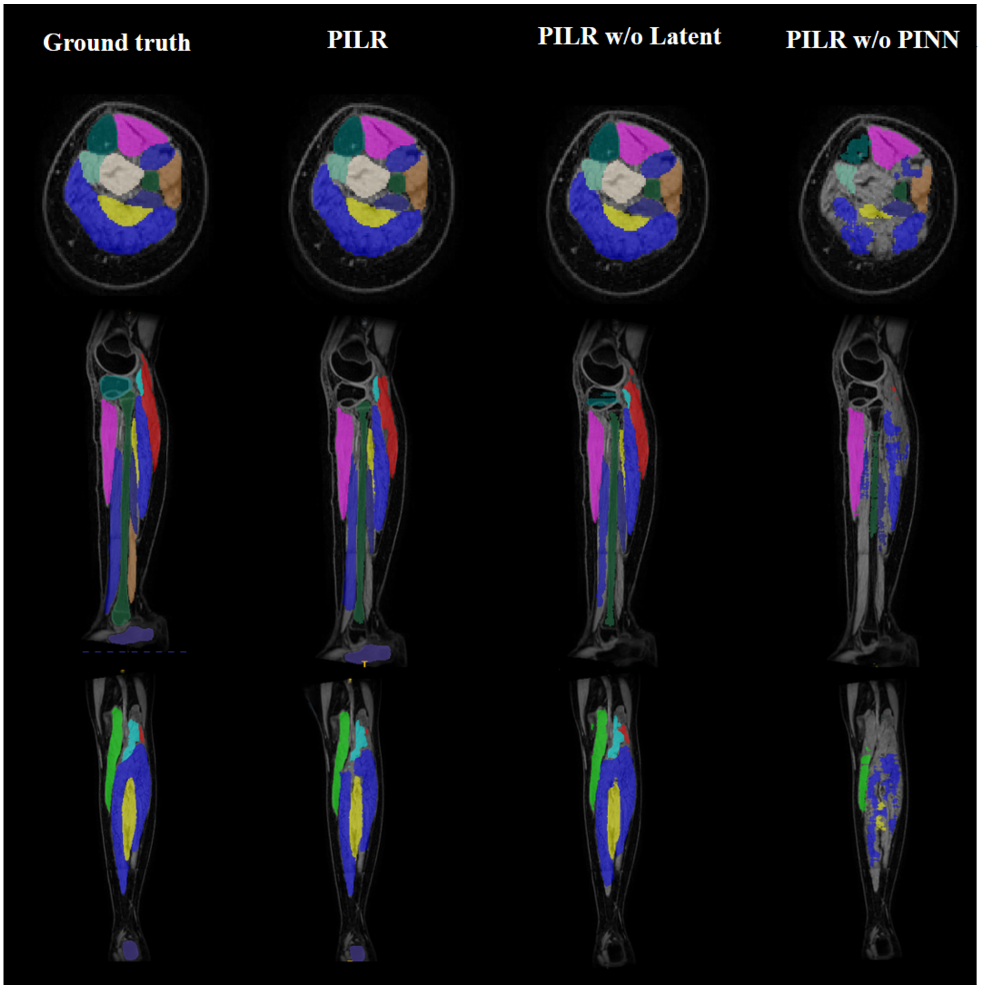


**Supplementary Figure (3).** Ablation study comparing the complete **PILR-U-Net** with two reduced variants: **PILR-U-Net (w/o Latent Regularization)** and **PILR-U-Net (w/o Physics-Informed Regularization) for four different subjects**. The model demonstrates close anatomical agreement with expert references, accurately preserving muscle boundaries and structural details.

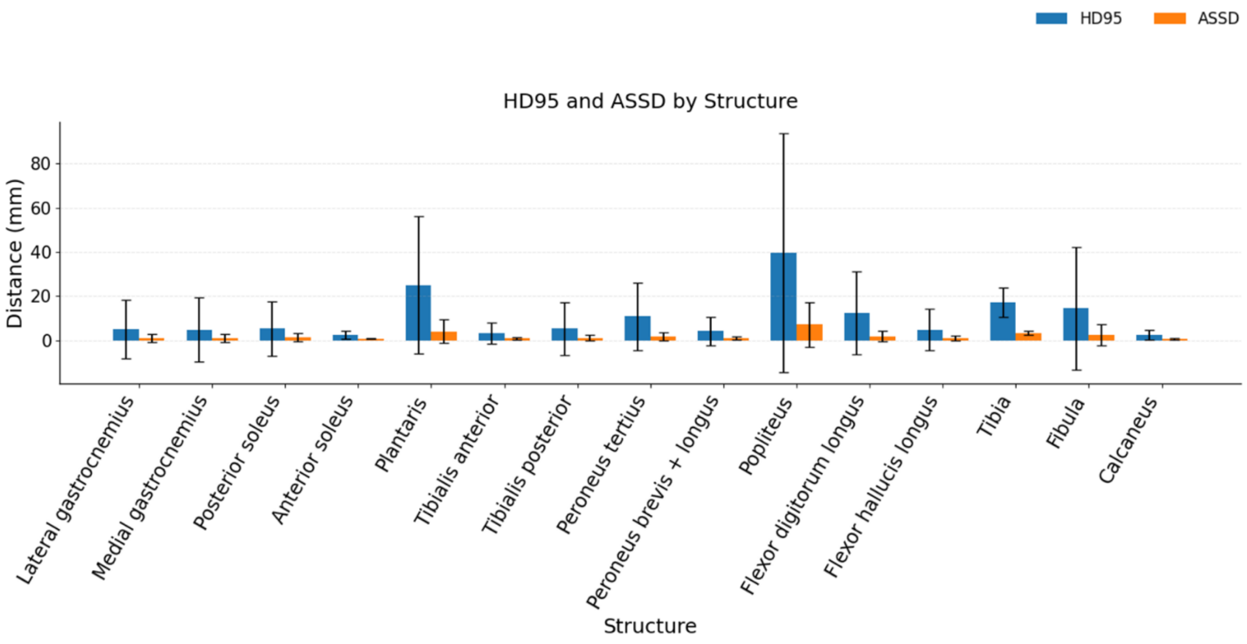


**Supplementary Figure 4.** Boundary accuracy (distance-based metrics) Boundary accuracy evaluated using the 95th percentile Hausdorff Distance (HD95) and Average Symmetric Surface Distance (ASSD). Lower values indicate improved boundary alignment between predictions and ground truth. While most structures exhibit low boundary errors, increased variability in smaller or anatomically complex muscles highlights the challenge of precise edge localization.

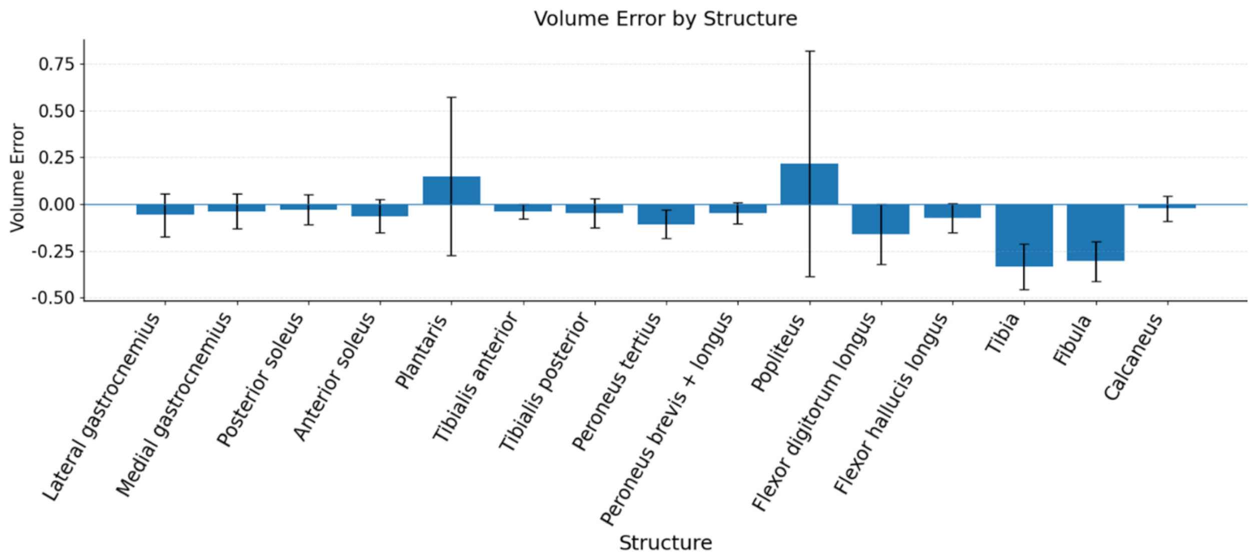


**Supplementary Figure 5.** (Volume estimation error) Volume error across anatomical structures, reflecting the relative difference between predicted and ground truth volumes. Values near zero indicate accurate volumetric reconstruction, while positive and negative deviations represent over- and under-segmentation, respectively. Overall, the model maintains low volumetric bias across the majority of structures.

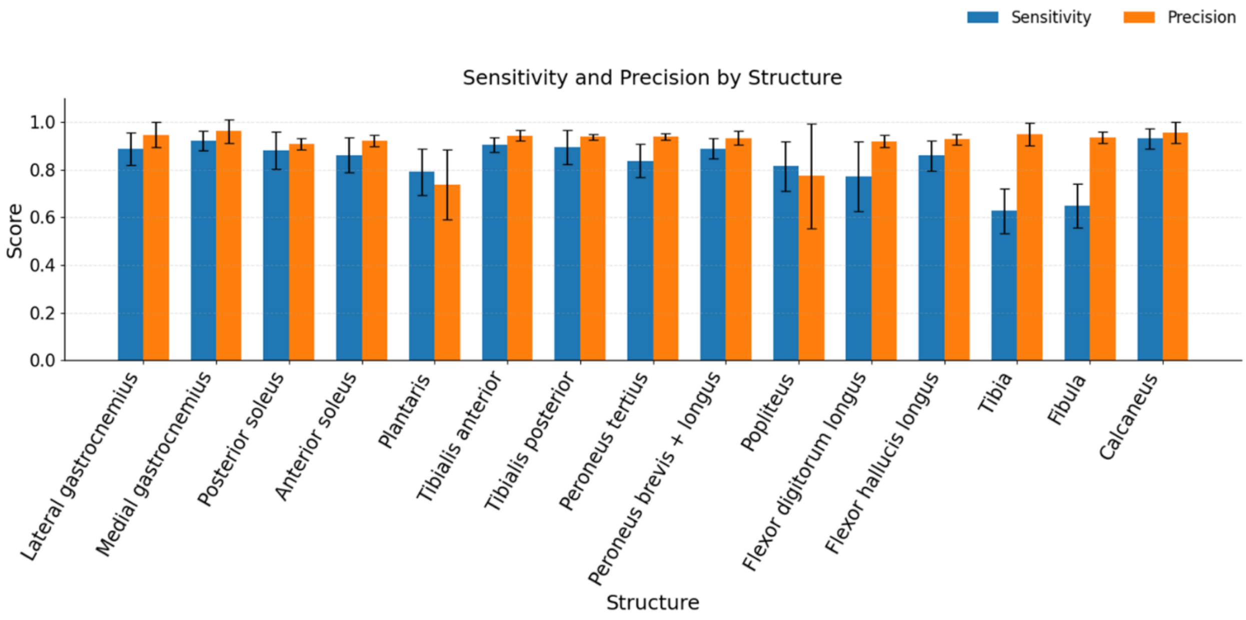


**Supplementary Figure 6.** Detection performance (sensitivity and precision) Sensitivity and precision metrics assessing voxel-wise detection performance. High sensitivity indicates effective identification of true positive regions, while high precision reflects low false positive rates. The balanced performance across both metrics demonstrates reliable segmentation with minimal over- and under-segmentation.

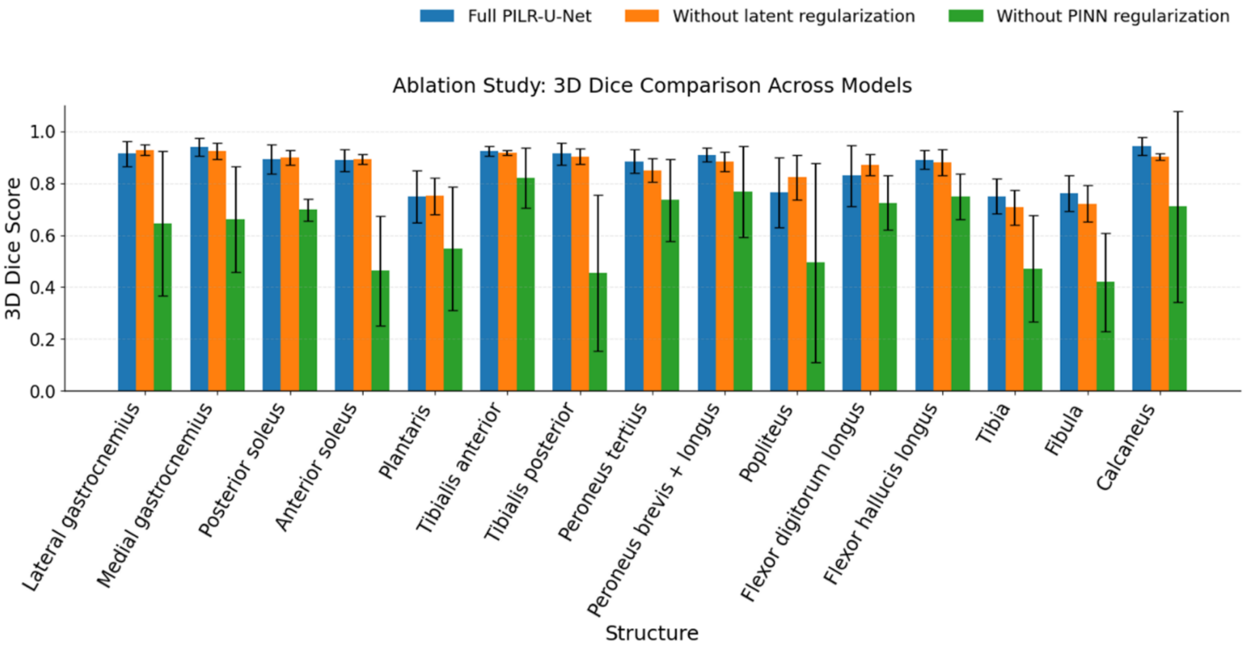


**Supplementary Figure 7.** Comparison of 3D Dice Similarity Coefficient across all evaluated muscles and bones for the complete PILR-U-Net framework, a variant without latent-space regularization, and a variant without physics-informed (PINN) regularization. Error bars represent one standard deviation across the test cohort. Removal of latent-space regularization resulted in moderate reductions in segmentation accuracy for several structures, whereas removal of the physics-informed regularization produced substantially larger decreases in Dice scores across nearly all muscles and bones. These findings demonstrate that both components contribute to segmentation performance, with the physics-informed regularization providing the largest overall benefit.

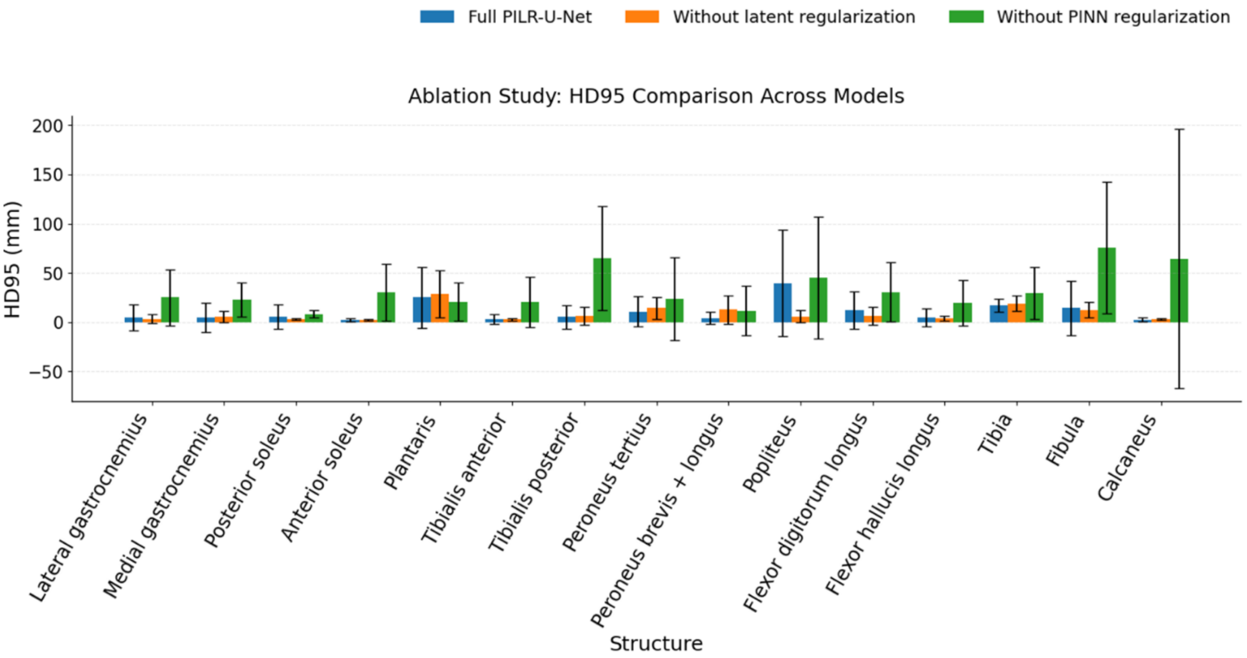


**Supplementary Figure 8.** Comparison of the 95th percentile Hausdorff Distance (HD95) across all evaluated muscles and bones for the complete PILR-U-Net framework, a variant without latent-space regularization, and a variant without physics-informed (PINN) regularization. Error bars represent one standard deviation across the test cohort. Removal of either regularization component increased boundary error; however, omission of the physics-informed regularization resulted in substantially larger HD95 values and greater variability, particularly for anatomically challenging structures such as the plantaris, popliteus, tibia, and fibula. These results indicate that the physics-informed constraints play a critical role in maintaining boundary accuracy and anatomical consistency.

| Structure | 3D Dice (Mean ± SD) | Jaccard (Mean ± SD) | 2D Dice (Mean ± SD) | HD95 (mm, Mean ± SD) | ASSD (mm, Mean ± SD) | Volume Error % (Mean ± SD) | Sensitivity (Mean ± SD) | Precision (Mean ± SD) |
|---|---|---|---|---|---|---|---|---|
| Lateral gastrocnemius | 0.9138 ± 0.0483 | 0.8445 ± 0.0737 | 0.8072 ± 0.0940 | 5.0257 ± 13.296 | 0.9357 ± 1.693 | -0.0580 ± 0.1145 | 0.8875 ± 0.0669 | 0.9471 ± 0.0529 |
| Medial gastrocnemius | 0.9398 ± 0.0347 | 0.8882 ± 0.0562 | 0.8432 ± 0.0756 | 4.6726 ± 14.524 | 0.8628 ± 1.857 | -0.0378 ± 0.0927 | 0.9215 ± 0.0422 | 0.9617 ± 0.0490 |
| Posterior soleus | 0.8928 ± 0.0563 | 0.8101 ± 0.0760 | 0.8082 ± 0.0896 | 5.3158 ± 12.373 | 1.1754 ± 1.825 | -0.0301 ± 0.0804 | 0.8813 ± 0.0790 | 0.9081 ± 0.0248 |
| Anterior soleus | 0.8883 ± 0.0411 | 0.8013 ± 0.0625 | 0.7915 ± 0.0571 | 2.4870 ± 1.759 | 0.6976 ± 0.268 | -0.0639 ± 0.0876 | 0.8614 ± 0.0722 | 0.9214 ± 0.0245 |
| Plantaris | 0.7489 ± 0.0993 | 0.6079 ± 0.1197 | 0.5342 ± 0.1426 | 24.9945 ± 31.037 | 3.9621 ± 5.336 | 0.1482 ± 0.4236 | 0.7913 ± 0.0966 | 0.7376 ± 0.1481 |
| Tibialis anterior | 0.9237 ± 0.0198 | 0.8589 ± 0.0333 | 0.7744 ± 0.0578 | 3.1502 ± 4.680 | 0.8013 ± 0.508 | -0.0396 ± 0.0375 | 0.9056 ± 0.0304 | 0.9433 ± 0.0211 |
| Tibialis posterior | 0.9133 ± 0.0413 | 0.8428 ± 0.0633 | 0.8068 ± 0.0721 | 5.2338 ± 12.045 | 1.0196 ± 1.365 | -0.0465 ± 0.0785 | 0.8936 ± 0.0707 | 0.9376 ± 0.0126 |
| peroneus tertius | 0.8838 ± 0.0453 | 0.7944 ± 0.0649 | 0.7809 ± 0.0644 | 10.7926 ± 15.237 | 1.6634 ± 1.831 | -0.1073 ± 0.0772 | 0.8380 ± 0.0710 | 0.9389 ± 0.0137 |
| Peroneus brevis + longus | 0.9095 ± 0.0271 | 0.8352 ± 0.0437 | 0.7955 ± 0.0608 | 4.1329 ± 6.405 | 0.9845 ± 0.732 | -0.0474 ± 0.0556 | 0.8883 ± 0.0425 | 0.9334 ± 0.0280 |
| Popliteus | 0.7646 ± 0.1354 | 0.6365 ± 0.1646 | 0.5342 ± 0.1925 | 39.6145 ± 54.048 | 7.0171 ± 10.206 | 0.2158 ± 0.6023 | 0.8153 ± 0.1034 | 0.7738 ± 0.2201 |
| Flexor digitorum longus | 0.8297 ± 0.1177 | 0.7217 ± 0.1315 | 0.6850 ± 0.1471 | 12.3769 ± 18.817 | 1.8214 ± 2.321 | -0.1609 ± 0.1607 | 0.7718 ± 0.1454 | 0.9199 ± 0.0266 |
| Flexor hallucis longus | 0.8905 ± 0.0352 | 0.8044 ± 0.0549 | 0.7769 ± 0.0726 | 4.7640 ± 9.342 | 1.0010 ± 1.029 | -0.0733 ± 0.0771 | 0.8590 ± 0.0633 | 0.9279 ± 0.0225 |
| Tibia | 0.7497 ± 0.0674 | 0.6041 ± 0.0854 | 0.7895 ± 0.0356 | 17.2072 ± 6.726 | 3.3228 ± 1.049 | -0.3344 ± 0.1204 | 0.6275 ± 0.0947 | 0.9484 ± 0.0487 |
| Fibula | 0.7610 ± 0.0681 | 0.6186 ± 0.0809 | 0.7979 ± 0.0773 | 14.4375 ± 27.807 | 2.3185 ± 4.872 | -0.3048 ± 0.1064 | 0.6484 ± 0.0912 | 0.9348 ± 0.0248 |
| Calcaneus | 0.9427 ± 0.0331 | 0.8933 ± 0.0572 | 0.8130 ± 0.0966 | 2.4462 ± 2.218 | 0.5911 ± 0.358 | -0.0243 ± 0.0685 | 0.9311 ± 0.0432 | 0.9566 ± 0.0451 |

**Supplementary Table 1.** Quantitative performance of the PILR-U-Net framework across 15 individual muscles. Metrics include volumetric overlap (3D Dice, Jaccard), slice-wise accuracy (2D Dice), boundary delineation (HD95, ASSD), volumetric consistency (Volume Error), and classification performance (Sensitivity, Precision). Values are reported as mean ± standard deviation, demonstrating robust segmentation accuracy and consistent inter-subject performance.

| Structure | 3D Dice (Mean ± SD) | Jaccard (Mean ± SD) | 2D Dice (Mean ± SD) | HD95 (mm, Mean ± SD) | ASSD (mm, Mean ± SD) | Volume Error % (Mean ± SD) | Sensitivity (Mean ± SD) | Precision (Mean ± SD) |
|---|---|---|---|---|---|---|---|---|
| Lateral gastrocnemius | 0.9283 ± 0.0195 | 0.8668 ± 0.0329 | 0.8032 ± 0.0777 | 3.3858 ± 4.6536 | 0.6940 ± 0.4953 | -3.3686 ± 4.5006 | 0.9130 ± 0.0364 | 0.9452 ± 0.0159 |
| Medial gastrocnemius | 0.9248 ± 0.0308 | 0.8617 ± 0.0516 | 0.7862 ± 0.1080 | 5.3394 ± 5.6517 | 0.9358 ± 0.6388 | -7.7232 ± 5.1899 | 0.8899 ± 0.0517 | 0.9643 ± 0.0112 |
| Posterior soleus | 0.8992 ± 0.0282 | 0.8181 ± 0.0459 | 0.8252 ± 0.0389 | 3.0943 ± 1.0843 | 0.8007 ± 0.2132 | 0.6062 ± 4.2316 | 0.9019 ± 0.0329 | 0.8973 ± 0.0340 |
| Anterior soleus | 0.8927 ± 0.0199 | 0.8067 ± 0.0322 | 0.7717 ± 0.0435 | 2.4037 ± 0.9860 | 0.6733 ± 0.1456 | -5.5655 ± 4.3491 | 0.8682 ± 0.0368 | 0.9196 ± 0.0122 |
| Plantaris | 0.7512 ± 0.0701 | 0.6066 ± 0.0907 | 0.4847 ± 0.1417 | 28.4171 ± 23.9676 | 4.2721 ± 3.2380 | 1.7352 ± 19.0599 | 0.7574 ± 0.0936 | 0.7562 ± 0.0894 |
| Tibialis anterior | 0.9176 ± 0.0101 | 0.8480 ± 0.0174 | 0.7509 ± 0.0486 | 2.9857 ± 1.2983 | 0.8069 ± 0.3097 | -2.8694 ± 3.4855 | 0.9046 ± 0.0226 | 0.9317 ± 0.0155 |
| Tibialis posterior | 0.9030 ± 0.0298 | 0.8245 ± 0.0474 | 0.7727 ± 0.0566 | 6.1939 ± 9.2896 | 1.2065 ± 0.9995 | -4.7301 ± 6.7589 | 0.8825 ± 0.0545 | 0.9272 ± 0.0214 |
| peroneus tertius | 0.8492 ± 0.0453 | 0.7406 ± 0.0658 | 0.7467 ± 0.0566 | 14.2988 ± 11.0713 | 2.0092 ± 1.0696 | -14.9947 ± 8.8429 | 0.7875 ± 0.0766 | 0.9273 ± 0.0149 |
| Peroneus brevis + longus | 0.8841 ± 0.0380 | 0.7943 ± 0.0586 | 0.7322 ± 0.0952 | 12.6905 ± 14.4512 | 1.8671 ± 1.4512 | -9.0447 ± 8.7853 | 0.8456 ± 0.0708 | 0.9312 ± 0.0223 |
| Popliteus | 0.8227 ± 0.0862 | 0.7073 ± 0.1168 | 0.6654 ± 0.0960 | 5.9407 ± 5.8714 | 1.2013 ± 0.8034 | -13.6620 ± 15.9971 | 0.7718 ± 0.1293 | 0.8985 ± 0.0520 |
| Flexor digitorum longus | 0.8720 ± 0.0407 | 0.7753 ± 0.0621 | 0.7274 ± 0.0716 | 6.3054 ± 8.8339 | 1.0784 ± 0.8308 | -10.2192 ± 7.6465 | 0.8289 ± 0.0687 | 0.9237 ± 0.0169 |
| Flexor hallucis longus | 0.8795 ± 0.0504 | 0.7883 ± 0.0747 | 0.7519 ± 0.0764 | 4.0848 ± 2.6296 | 0.8912 ± 0.4028 | -8.7896 ± 7.5106 | 0.8426 ± 0.0769 | 0.9231 ± 0.0233 |
| Tibia | 0.7073 ± 0.0665 | 0.5513 ± 0.0798 | 0.7681 ± 0.0463 | 18.9067 ± 7.7336 | 3.6967 ± 1.0128 | -33.2905 ± 19.2651 | 0.5947 ± 0.1178 | 0.9125 ± 0.0812 |
| Fibula | 0.7215 ± 0.0712 | 0.5692 ± 0.0865 | 0.7814 ± 0.0400 | 12.3773 ± 7.9015 | 1.8866 ± 0.8672 | -36.1935 ± 10.9206 | 0.5947 ± 0.0964 | 0.9339 ± 0.0236 |
| Calcaneus | 0.9027 ± 0.0125 | 0.8229 ± 0.0207 | 0.7263 ± 0.0071 | 3.0310 ± 1.0310 | 0.8828 ± 0.2312 | -4.5726 ± 9.7104 | 0.8827 ± 0.0560 | 0.9286 ± 0.0358 |

**Supplementary Table 2.** Quantitative performance of the proposed framework without latent regularization across 15 individual muscles. Metrics include volumetric overlap (3D Dice, Jaccard), slice-wise accuracy (2D Dice), boundary delineation (HD95, ASSD), volumetric consistency (Volume Error), and classification performance (Sensitivity, Precision). Values are reported as mean ± standard deviation, demonstrating robust segmentation accuracy and consistent inter-subject performance.

| Structure | 3D Dice (Mean ± SD) | Jaccard (Mean ± SD) | 2D Dice (Mean ± SD) | HD95 (mm, Mean ± SD) | ASSD (mm, Mean ± SD) | Volume Error % (Mean ± SD) | Sensitivity (Mean ± SD) | Precision (Mean ± SD) |
|---|---|---|---|---|---|---|---|---|
| Lateral gastrocnemius | 0.6452 ± 0.2797 | 0.5328 ± 0.2773 | 0.5228 ± 0.2593 | 25.1398 ± 28.3781 | 4.8351 ± 5.5439 | -35.3632 ± 35.5970 | 0.5782 ± 0.3082 | 0.9144 ± 0.0653 |
| Medial gastrocnemius | 0.6620 ± 0.2031 | 0.5263 ± 0.2101 | 0.5052 ± 0.2065 | 22.9010 ± 17.0974 | 4.1023 ± 2.8113 | -42.9164 ± 22.8446 | 0.5429 ± 0.2188 | 0.9517 ± 0.0227 |
| Posterior soleus | 0.6976 ± 0.0432 | 0.5374 ± 0.0514 | 0.6122 ± 0.0734 | 8.3605 ± 4.0251 | 2.3858 ± 0.6919 | -14.6619 ± 11.6270 | 0.6468 ± 0.0589 | 0.7654 ± 0.0728 |
| Anterior soleus | 0.4633 ± 0.2119 | 0.3244 ± 0.1673 | 0.3314 ± 0.1734 | 30.4426 ± 28.9854 | 6.6166 ± 5.8622 | -48.0423 ± 33.5117 | 0.3817 ± 0.2221 | 0.7229 ± 0.2767 |
| Plantaris | 0.5483 ± 0.2376 | 0.4106 ± 0.2037 | 0.4086 ± 0.2227 | 20.6867 ± 19.3169 | 4.5011 ± 4.8719 | -43.8718 ± 30.9666 | 0.4631 ± 0.2455 | 0.8056 ± 0.0944 |
| Tibialis anterior | 0.8208 ± 0.1155 | 0.7109 ± 0.1510 | 0.6019 ± 0.1636 | 20.6031 ± 25.5765 | 3.2608 ± 3.1176 | -21.4168 ± 16.2121 | 0.7422 ± 0.1586 | 0.9428 ± 0.0183 |
| Tibialis posterior | 0.4546 ± 0.3009 | 0.3434 ± 0.2547 | 0.3642 ± 0.2677 | 64.9468 ± 53.1605 | 12.7782 ± 12.0922 | -59.5548 ± 29.5371 | 0.3625 ± 0.2740 | 0.8385 ± 0.1890 |
| peroneus tertius | 0.7348 ± 0.1568 | 0.6018 ± 0.1703 | 0.6462 ± 0.1819 | 23.4558 ± 42.1849 | 4.4759 ± 7.5307 | -29.4692 ± 17.8465 | 0.6404 ± 0.1798 | 0.8974 ± 0.0484 |
| Peroneus brevis + longus | 0.7666 ± 0.1748 | 0.6490 ± 0.1958 | 0.6607 ± 0.1833 | 11.7222 ± 24.9176 | 2.2534 ± 2.9309 | -21.9473 ± 19.3990 | 0.6987 ± 0.2066 | 0.8778 ± 0.0945 |
| Popliteus | 0.4941 ± 0.3824 | 0.4149 ± 0.3441 | 0.3872 ± 0.3263 | 45.2291 ± 61.9566 | 30.4787 ± 50.3124 | -36.1965 ± 34.2723 | 0.4425 ± 0.3668 | 0.6072 ± 0.4075 |
| Flexor digitorum longus | 0.7240 ± 0.1049 | 0.5772 ± 0.1194 | 0.5626 ± 0.1502 | 30.5644 ± 30.2729 | 4.4291 ± 4.2802 | -27.7606 ± 20.5215 | 0.6334 ± 0.1525 | 0.8900 ± 0.0538 |
| Flexor hallucis longus | 0.7483 ± 0.0867 | 0.6053 ± 0.1066 | 0.5621 ± 0.1258 | 19.5380 ± 23.3123 | 3.4025 ± 3.0386 | -20.7180 ± 16.1972 | 0.6762 ± 0.1251 | 0.8599 ± 0.0677 |
| Tibia | 0.4717 ± 0.2059 | 0.3319 ± 0.1739 | 0.5638 ± 0.1965 | 29.6210 ± 26.2362 | 7.4469 ± 6.6718 | -62.8270 ± 20.9461 | 0.3445 ± 0.1862 | 0.9317 ± 0.0608 |
| Fibula | 0.4196 ± 0.1888 | 0.2840 ± 0.1557 | 0.4515 ± 0.2078 | 75.8187 ± 66.6984 | 13.5313 ± 12.5400 | -63.5058 ± 23.1446 | 0.3072 ± 0.1830 | 0.8501 ± 0.0538 |
| Calcaneus | 0.7108 ± 0.3677 | 0.6507 ± 0.3514 | 0.6340 ± 0.3521 | 64.5338 ± 131.8120 | 51.2493 ± 116.1249 | 16.7137 ± 67.5719 | 0.6840 ± 0.3547 | 0.7407 ± 0.3833 |

**Supplementary Table 3.** Quantitative performance of the proposed framework without PINN regularization across 15 individual muscles. Metrics include volumetric overlap (3D Dice, Jaccard), slice-wise accuracy (2D Dice), boundary delineation (HD95, ASSD), volumetric consistency (Volume Error), and classification performance (Sensitivity, Precision). Values are reported as mean ± standard deviation, demonstrating robust segmentation accuracy and consistent inter-subject performance.

| Structure | Full mean | No latent mean | No PINN mean | Friedman p FDR | Full vs No latent p FDR | Full vs No PINN p FDR |
|---|---|---|---|---|---|---|
| Lateral gastrocnemius | 0.923833333 | 0.927367845 | 0.645201207 | 0.005576609 | 0.802283654 | 0.006510417 |
| Medial gastrocnemius | 0.93483 | 0.924831142 | 0.661951577 | 0.001659253 | 0.690569196 | 0.004882813 |
| Posterior soleus | 0.87285 | 0.899221119 | 0.697646875 | 0.001659253 | 0.263671875 | 0.004882813 |
| Anterior soleus | 0.88422 | 0.892665834 | 0.41697327 | 0.001659253 | 0.802283654 | 0.004882813 |
| Plantaris | 0.7872375 | 0.768972331 | 0.548256752 | 0.099658381 | 0.802283654 | 0.063100962 |
| Tibialis anterior | 0.91416 | 0.917630729 | 0.820840587 | 0.011222382 | 0.824497768 | 0.006510417 |
| Tibialis posterior | 0.88719 | 0.903028405 | 0.454643427 | 0.001659253 | 0.845703125 | 0.004882813 |
| Peroneus tertius | 0.84016 | 0.849248397 | 0.73481673 | 0.077544822 | 0.802283654 | 0.16015625 |
| Peroneus brevis + longus | 0.89997 | 0.884101111 | 0.766550193 | 0.005576609 | 0.251953125 | 0.013316761 |
| Popliteus | 0.78465 | 0.822691362 | 0.494060727 | 0.122456428 | 0.802283654 | 0.089983259 |
| Flexor digitorum longus | 0.82851 | 0.87202749 | 0.724018001 | 0.001688847 | 0.241699219 | 0.013316761 |
| Flexor hallucis longus | 0.8837 | 0.879500295 | 0.74834996 | 0.011169875 | 0.802283654 | 0.006510417 |
| Tibia | 0.76122 | 0.707302929 | 0.471709391 | 0.002637668 | 0.241699219 | 0.004882813 |
| Fibula | 0.75965 | 0.721519399 | 0.419554673 | 0.001659253 | 0.241699219 | 0.004882813 |
| Calcaneus | 0.933222222 | 0.302280656 | 0.683811714 | 0.039903115 | 0.17578125 | 0.034179688 |

**Supplementary Table 4. Statistical analysis of the ablation study based on the mean 3D Dice coefficient for each musculoskeletal structure.** Mean segmentation performance is reported for the complete **PILR-U-Net**, **PILR-U-Net without latent regularization**, and **PILR-U-Net without physics-informed regularization**. Overall differences among the three model variants were assessed using the Friedman test, followed by pairwise Wilcoxon signed-rank tests comparing the complete model with each ablation model. Reported p-values were corrected for multiple comparisons using the Benjamini–Hochberg false discovery rate (FDR) procedure. Statistically significant differences (FDR-corrected $p < 0.05$) indicate that removal of the corresponding regularization component significantly affected segmentation performance.

| Structure | Full mean | No latent mean | No PINN mean | Friedman p FDR | Full vs No latent p FDR | Full vs No PINN p FDR |
|---|---|---|---|---|---|---|
| Lateral gastrocnemius | 2.677111111 | 3.586332869 | 25.13982487 | 0.008880518 | 0.719401042 | 0.014648438 |
| Medial gastrocnemius | 3.5097 | 5.33936658 | 22.90099072 | 0.046356612 | 0.654296875 | 0.051269531 |
| Posterior soleus | 5.7055 | 3.094254477 | 8.360540642 | 0.018616458 | 0.719401042 | 0.048828125 |
| Anterior soleus | 2.1835 | 2.403690129 | 39.72632698 | 0.002780577 | 0.719401042 | 0.009765625 |
| Plantaris | 14.091375 | 24.13216293 | 20.68667864 | 0.246139594 | 0.719401042 | 0.576171875 |
| Tibialis anterior | 6.4881 | 2.98571425 | 20.60314514 | 0.046356612 | 0.921875 | 0.029296875 |
| Tibialis posterior | 11.7367 | 6.193875455 | 64.94683679 | 0.002780577 | 0.721153846 | 0.009765625 |
| Peroneus tertius | 22.1727 | 14.29882365 | 23.45575686 | 0.322708084 | 0.744977679 | 0.921875 |
| Peroneus brevis + longus | 5.8624 | 12.69048968 | 11.72224557 | 0.018616458 | 0.146484375 | 0.051269531 |
| Popliteus | 31.3702 | 5.940696915 | 45.22912292 | 0.314459761 | 0.719401042 | 0.887920673 |
| Flexor digitorum longus | 18.2425 | 6.305437138 | 30.56438705 | 0.904837418 | 0.721153846 | 0.906110491 |
| Flexor hallucis longus | 4.3045 | 4.08478744 | 19.53799997 | 0.067573804 | 0.719401042 | 0.061848958 |
| Tibia | 16.4504 | 18.90674496 | 29.62100986 | 0.203957208 | 0.721153846 | 0.096679688 |
| Fibula | 11.2089 | 12.37734887 | 75.81868156 | 0.018616458 | 0.721153846 | 0.009765625 |
| Calcaneus | 3.257888889 | 221.1094583 | 71.56817918 | 0.053204153 | 0.146484375 | 0.17578125 |

**Supplementary Table 5. Statistical analysis of the ablation study based on the mean 95th percentile Hausdorff Distance (HD95) for each musculoskeletal structure.** Mean HD95 values (mm) are reported for the complete **PILR-U-Net**, **PILR-U-Net without latent regularization**, and **PILR-U-Net without physics-informed regularization**. Lower HD95 values indicate improved boundary delineation and greater agreement with the reference segmentations. Overall differences among the three model variants were assessed using the Friedman test, followed by pairwise Wilcoxon signed-rank tests comparing the complete model with each ablation model. Reported *p*-values were corrected for multiple comparisons using the Benjamini–Hochberg false discovery rate (FDR) procedure. Statistically significant differences (FDR-corrected $p < 0.05$) indicate that removal of the corresponding regularization component significantly affected boundary accuracy.
